# Experiential acceptance during an episode of anxiety: Conceptualizing the process of acceptance through a qualitative study

**DOI:** 10.64898/2026.04.03.26346604

**Authors:** Julie Ribeyron, Nathalie Duriez, Rebecca Shankland

## Abstract

**Introduction:** Experiential acceptance refers to the capacity to be open to internal experiences without attempting to change or avoid them. Although acceptance is a core emotion regulation strategy within mindfulness- and acceptance-based interventions (MABIs) and a protective factor for mental health, its conceptualization and implementation remain unclear and ambiguous. The aim of this study was to clarify and develop a comprehensive model of accepting anxiety.

**Method:** Twenty-six participants from a non-clinical sample with prior experience in MABIs took part in semi-structured interviews exploring their experience of accepting anxiety. Data collection and analysis followed the principles of Grounded Theory to generate a data-driven model of the acceptance process.

**Results:** We identified a five-stage dynamic model involving distinct processes: (Stage 1) observing through the body with attentional focus on interoceptive experience; (Stage 2) identifying and acknowledging anxiety; (Stage 3) validating and normalizing the experience through validation and self-compassion; (Stage 4) not reacting characterized by decentering and nonreactivity; and (Stage 5) staying with the experience via exposure. We also identified facilitating factors that support engagement in the acceptance process.

**Conclusion:** These findings refine the understanding of acceptance as a multidimensional emotion regulation process by highlighting an active dynamic involving multiple mechanisms underlying the acceptance of anxiety. This model provides a framework for developing more targeted clinical interventions and for investigating individual and contextual variability in these subprocesses.

## Introduction

Clinical psychology has progressively shifted toward a transdiagnostic, process-based perspective that emphasizes underlying psychological processes shared across disorders, rather than diagnostic categories derived from the medical model (1). Processes were initially defined as cognitive or behavioral aspects that maintain psychological disorder (2) and has since been expanded to encompass metacognitive, cognitive, emotional, behavioral, and motivational processes, viewed as dynamic and transformational (3). More recently, therapeutic processes have been conceptualized as theory-driven, dynamic, and multi-level changes unfolding in empirically supported sequences that produce clinical benefits (4). This perspective supports integrative models and clinical interventions targeting common processes, notably emotion regulation (5). Emotion regulation is the activation of a goal to change an emotional trajectory (6). It plays a key role on the etiology and maintenance of psychopathology, particularly emotional disorders, (7). Emotion regulation can be implemented through a range of strategies, among which acceptance is a particularly relevant target because it is typically classified as both adaptive (8) and engagement-based, reflecting an orientation toward, rather than away from, emotional experience (9), and is consistently associated with better mental health outcomes, including lower psychopathology (e.g., 10–12).

Acceptance refers to experiential openness to present-moment emotional experience, pleasant or unpleasant, without attempts to avoid it (13), and involves a conscious decision to relinquish efforts to have different internal experiences, alongside an active willingness to feel emotions, observe thoughts, and allow memories to arise as they are (14). Unlike many emotion regulation strategies, acceptance occupies a distinctive position (8,9), because it does not aim to directly modify emotional intensity or quality, as emphasized in traditional definitions of emotion regulation (6), but instead involves allowing emotions to be present as they are and changing how individuals relate to them.

### The central role of acceptance in mindfulness- and acceptance-based interventions

Acceptance lies at the core of mindfulness and acceptance-based interventions (MABIs). These approaches include mindfulness-based interventions (MBI), such as Mindfulness-Based Stress Reduction (MBSR; 15) and Mindfulness-Based Cognitive Therapy (MBCT; 16), as well as Acceptance and Commitment Therapy (ACT; 14), and Dialectical Behavior Therapy (DBT; 17). Dismantling studies have provided converging evidence that acceptance is a central mechanism underlying emotion regulation and stress reduction in MBI (18), whereas attentional monitoring alone, without acceptance, may heighten awareness of negative emotional states and increase affective reactivity. In contrast, acceptance appears to reduce reactivity by altering the individual’s relationship to internal experience (19). Consistently, a meta-analysis (20) concluded that acceptance is the active component of MBI, while highlighting the need for clearer conceptualization and operationalization of acceptance. Acceptance has been most extensively examined within mindfulness theories and interventions. Although early work primarily established the efficacy of MABIs, research has increasingly focused on mechanisms of change, with acceptance commonly framed as both a core component of mindfulness and a key mechanism. Bishop et al. (21) conceptualized mindfulness as comprising attention monitoring and an accepting orientation toward experience. Other theoretical models have sought to articulate the mechanisms underlying mindfulness, either explicitly or implicitly incorporating acceptance. For instance, Baer (22) identified acceptance alongside exposure, cognitive change, self-regulation, and relaxation as central mechanisms underlying mindfulness-based practices. Similarly, Shapiro et al. (23), identified attention, intention (i.e., the motivation for practice), and attitude (i.e., the manner in which attention is deployed, characterized by patience, compassion, and non-attachment rather than struggle) as the core components of mindfulness, summarized in the formulation *“paying attention on purpose, in a particular way.”* with acceptance primarily embedded in attitude as an open, curious stance and, secondarily, in intention. Brown et al. (24) further proposed that mindfulness benefits are mediated by processes such as insight (i.e., a decentered perspective on transient thoughts and emotion) and voluntary exposure, which reduces emotional reactivity by facilitating contact with unpleasant experiences, with nonattachment supporting willingness and present-moment choice. Drawing on Buddhist psychology, Grabovac et al. (25) model identifies acceptance and compassion among key processes, including, conceptualizing them as attitudinal qualities that facilitate attentional refocusing by limiting judgment and discursive thought, and framing acceptance as a “quality of awareness” rather than a cognitive operation. A further conceptualization emerged from psychometric research on the Five Facets Mindfulness Questionnaire (FFMQ, 26), which identifies five dimensions: observing, describing, acting with awareness, nonjudging of inner experience, and nonreactivity to inner experience. However, it remains unclear to what extent the FFMQ adequately captures acceptance. While some studies operationalize acceptance primarily through the nonjudging facet (27), others include both nonjudging and nonreactivity (28), a position more consistent with the original conceptualization. Although not explicitly foregrounding acceptance, Hölzel et al. (29) identified attentional regulation, body awareness, emotion regulation and change in perspective on the self as core mindfulness mechanisms, with emotion regulation linked to exposure with response prevention (ERP; 30), whereby repeated, voluntary contact with feared internal or external cues in the absence of defensive responses diminishes conditioned reactions through new learning (31,32) as well as to memory reconsolidation accounts (33,34). Finally, some models emphasize attentional regulation as primary (35–37), viewing acceptance either as an antecedent stance facilitating attentional focus (25) or as a consequence of attentional training (38). Acceptance is also a core ACT construct and a key process underpinning psychological flexibility (14,39) defined as the capacity to remain in contact with the present moment and to persist in or change behavior to support personally meaningful goals (40). Acceptance is one of six core processes in ACT hexaflex model alongside cognitive defusion, self-as-context, contact with the present moment, values, and committed action. It primarily functions to facilitate values-based action, rather than constituting a therapeutic goal in itself (14,40).

#### Effects of acceptance on anxiety

Research on emotion regulation has highlighted the importance of examining specific emotions, as strategies may vary depending on the emotional target and anxiety is therefore a particularly relevant focus. Anxiety is an emotion characterized by apprehension and somatic tension (e.g., muscle tension, faster breathing, increased heart rate), reflecting anticipation of potential danger, catastrophe, or misfortune. Anxiety is typically a future-oriented, long-acting response to a diffuse perceived threat, whereas fear is a brief, present-oriented response triggered by an identifiable threat (45). Like other emotions, anxiety typically involves transient responses. However, when these reactions become persistent, intense, and intrusive, they may reflect pathological anxiety or an emotional disorder. Anxiety disorders are classified in the DSM as symptom-based categories, including generalized anxiety disorder, phobias, and panic disorder, among others. Importantly, anxiety is not limited to these diagnostic categories but is also present across a wide range of mental and somatic conditions. Systematic reviews have demonstrated the beneficial effects of MABIs in reducing anxiety symptoms (46,47), including when such interventions are delivered online (48).

### Moderators related to acceptance

Factors influencing emotion regulation remain relatively understudied when it comes specifically to acceptance. However, self-compassion has been identified as a key factor associated with the use of adaptive emotion regulation strategies (49,50), including acceptance. Self-compassion (51) refers to a kind and supportive attitude toward oneself in the face of suffering and comprises self-kindness (i.e., responding to personal difficulties with warmth and understanding rather than self-criticism), common humanity (i.e., recognizing one’s difficulties as part of the shared human experience) and mindfulness (i.e., balanced awareness of emotions without avoidance or exaggeration). Self-compassion has been consistently associated with more effective emotion regulation, lower levels of anxiety and depression, and enhanced psychological well-being (52).

### Aims of the study

Most research on acceptance has relied on quantitative methods, primarily self-report measures, that capture outcomes more than lived process. For this reason, qualitative approaches have been recommended to better capture the underlying processes of change (54), including how acceptance is enacted in practice, its stages, the difficulties encountered, the factors that moderate its effectiveness, and the perceived benefits beyond symptom reduction (55). Although existing findings suggest that accepting anxiety is beneficial, several questions remain insufficiently explored: How do individuals actually operationalize acceptance of their anxiety in everyday life? Which factors influence their capacity to engage in acceptance? What consequences do they perceive for their anxiety experience and broader emotion regulation? To address these questions, the present study aims to examine how acceptance of anxiety is implemented in practice by identifying its stages, underlying mechanisms, and constituent processes. More specifically, the objectives are to (a) describe the acceptance process as it is subjectively experienced, (b) identify factors that facilitate or hinder its implementation, and (c) explore perceived benefits extending beyond symptom reduction.

## Method

The study protocol was approved by the local ethics committee of the university where the study was conducted (protocol #CE-P8-2021-07). All participants provided written informed consent prior to data collection. This study was conducted in concordance with ethical standards, including the principals of the 1989 revised Helsinki Declaration and the American Psychology Association guidelines, regarding respect for data confidentiality, anonymization, free and informed consent, and the right of participants to withdraw from the research at any time.

### Participants

Recruitment and data collection took place between 2 September 2024 and 21 January 2025. Participants (N = 26) were recruited through an announcement on the professional social network LinkedIn, as well as via a professional association of MBSR mindfulness instructors and an association of ACT therapists. Eligibility criteria included being a French speaker, having completed at least one MABI delivered by a certified practitioner, and regularly using acceptance to cope with experiences of anxiety. Consistent with the principles of Grounded Theory, the sampling strategy evolved during the course of the study toward the recruitment of more experienced participants, many of whom were themselves instructors or therapists delivering MABIs. To ensure a broad range of perspectives and support theoretical saturation, both participants with limited experience and those with extensive professional experience were included in the interviews. Potential participants were invited to contact one of the co-authors via email. They received an information sheet describing the study and participation requirements, as well as an informed consent form to be signed prior to participation. A videoconference interview was then scheduled.

The final sample comprised 26 participants (7 men, 19 women), aged 29-68 years. Table 1 provided details on demographics, training backgrounds, years of practice, and instructor/therapist experience.

**Table 1.** Descriptive characteristics of the participants.

| N° | Gender | Age | Occupation | Primary MABI | Personal Practice (years) | Instructor/ACT Therapist experience (years) |
| --- | --- | --- | --- | --- | --- | --- |
| 1 | M | 20-29 | Psychologist | MBCT, ACT | 4, 2 | - |
| 2 | W | 40-49 | Sport teacher | Mindful'up, MBSR | 10 | - |
| 3 | W | 30-39 | Special Education Educator | MBSR | 4 | - |
| 4 | W | 50-59 | Psychologist | MBSR, Yoga | 4 | - |
| 5 | W | 30-39 | Special Education Teacher | MBCL, MBSR | 1 | - |
| 6 | W | 40-49 | Psychologist | MBCT, Fovea, Compassion | 16 | 10 |
| 7 | W | 30-39 | Psychologist | MBCT, Compassion, ACT | 8 | 5 |
| 8 | M | 50-59 | Psychologist | MBCT, Fovea | 15 | 11 |
| 9 | W | 50-59 | Psychologist | MBCT | 10 | 5 |
| 10 | W | 50-59 | Sophrologist, yoga teacher | MBSR, Yoga | 7 | 3 |
| 11 | W | 50-59 | Nurse | MBSR | 10 | 3 |
| 12 | W | 50-59 | Mindfulness instructor | MBSR | 20 | 6 |
| 13 | W | 40-49 | Psychologist | Compassion | 27 | 11 |
| 14 | W | 50-59 | Mindfulness instructor | MBSR | 20 | 5 |
| 15 | W | 50-59 | Coach | MBSR | 9 | 6 |
| 16 | W | 30-39 | Social Care Manager | MBSR | 5 | - |
| 17 | M | 60-69 | Osteopath (retired) | MBSR, Buddhism | 28 | 9 |
| 18 | W | 50-59 | Speech-Language Pathologist | ACT | 10 | 10 |
| 19 | W | 40-49 | Psychotherapy practitioner | ACT | 3 | 3 |
| 20 | M | 50-59 | Psychomotor Therapist | ACT | 2 | - |
| 21 | W | 30-39 | Psychologist | ACT, DBT | 3 | 3 |
| 22 | M | 60-69 | Peer Health Worker | ACT | 1 | - |
| 23 | W | 40-49 | Psychologist | ACT | 1 | 1 |
| 24 | M | 50-59 | Psychiatrist | ACT | 10 | 10 |
| 25 | M | 40-49 | Psychologist | ACT | 9 | 8 |
| 26 | W | 40-49 | Psychologist | ACT, MBCT | 10 | 10 |

Notes. M= man ; W= woman. MABI = mindfulness- and acceptance-based interventions; MBI = mindfulness-based interventions. MBCT = mindfulness-based cognitive therapy; MBSR = mindfulness-based stress reduction; MBCL = mindfulness-based compassionate living; ACT = acceptance and commitment therapy; DBT = dialectical behavior therapy. Practice (in years) refers to the number of years of personal practice in the main MABIs reported and/or practicing as an ACT therapist. A dash (–) indicates that the participant did not report instructor/therapist experience.

### Procedure

#### Data collection

Data were collected via videoconference using in-depth semi-structured interviews conducted by a co-author, included 19 open-ended questions covering six thematic areas (anxiety experience, typical emotion regulation strategies, acceptance understanding and practice, influencing factors, perceived effect). Interviews were audio-recorded and transcribed for analysis.

#### Data analysis

In line with Grounded Theory methodology (56), data collection and analysis were conducted concurrently, with each interview being collected, coded, and analyzed sequentially. Consistent with theoretical sampling, the interview guide was iteratively refined throughout the analytic process to address emerging gaps and needs identified in the data, allowing for flexibility in data collection to support theory development. Recruitment continued until theoretical saturation (i.e., no new concepts, properties, or dimensions emerged). The sample size was not predetermined; as analysis progressed, recruitment increasingly focused on more experienced MABIs practitioners to obtain richer practice-based data grounded and was extended to ACT therapists to broaden representation. Analytic memos were written throughout the process to examine relationships between codes and categories. The interviews were automatically transcribed and coded by one co-author using MAXQDA (57), with regular discussion with the co-authors. Analysis followed open, axial, and selective coding outlined in Grounded Theory and was guided by the constant comparative method to develop the analytic framework. Line-by-line open coding was first conducted to identify emerging concepts, which were subsequently grouped into eleven more abstract categories organized into three overarching axes. Relationships within and between categories were progressively identified through axial and selective coding. Finally, all transcripts were reread, and interviews with ACT practitioners were selectively recoded to further refine the analytic framework.

## Results

Rather than identifying a core category, the data supported a dynamic, temporally structured process-oriented model of accepting anxiety, spanning initial mobilization, the acceptance process, and subsequent perceived transformations. Accordingly, results are organized into three axes: (Axis 1) Choosing acceptance which examines obstacles to and facilitators of engaging in acceptance during anxiety; (Axis 2) Acceptance processes which outlines the successive stages on the acceptance process; and (Axis 3) Acceptance-related outcomes which identifies perceived effects on the anxiety experience and self-perception.

### Axis 1 – Deliberate engagement in acceptance

#### Anxiety as an intrusive and limiting emotional experience

> *“Sometimes, I’d feel completely overwhelmed—like I couldn’t do anything anymore or think clearly about anything at all.” (P23)*

Anxiety was described as intrusive and constraining, marked by repetitive thoughts *“getting stuck in my thoughts” (P3*) and cognitive overload *“everything feels kind of blurry in my head, it’s all over the place” (P1)*, which captured attention and made it difficult to remain anchored in the present moment. When anxiety intensifies, it could overwhelm the entire experiential field, *“it spills over” (P5),* making regulation difficult and acceptance even harder to consider. In some cases, the sense of intrusion was so pronounced that engaging in acceptance did not even seem like an option. Fatigue, accumulation of anxiety-provoking situations and links to past or early traumatic experiences further reduced access to acceptance.

#### Deliberately engaging in acceptance process

> *“It’s really about taking a second—disconnecting for a moment so I can actually get the whole thing started.” (P16)*

Recognizing the need for acceptance often difficult and effortful, particularly when the overwhelming nature of anxiety prevented the psychological distance necessary to access it. Because the default response was to resist or avoid anxiety, engaging in acceptance required a deliberate, proactive choice and sufficient mental or temporal availability to slow down and attend to the present experience: *“I think it really does require a daily practice, but also a certain kind of availability” (P12).* Several factors facilitated engagement in acceptance, including the MABI framework (e.g., meditation practices, metaphors), affective science and emotion-focused psychoeducation (i.e., understanding the nature and function of emotions and thoughts, and the limits of avoidance), reconnecting with meaning and personal values *“I often ask myself: what do I really want?” (P24)* as well as self-kindness and self-compassion *“It was really the self-compassion work that made that barrier finally break down” (P26)*, and prior experiences, personal or observed in others*“seeing over and over again that it works” (P24)*. However, choosing not to engage in acceptance and instead relying on another emotion regulation strategy can also be an adaptive response. When anxiety is too intense, initial downregulation may be needed, with or without later engaging in acceptance. From a self-compassionate perspective, this involves staying attuned to one’s needs and limits, in order not to maintain or amplify distress beyond what is tolerable: “*It’s not about this samurai stance, like no matter what, I stay in the storm. No—at some point, you have to take care of yourself, you have to move back and forth between exploring what’s there and what’s actually tolerable” (P15)*. More specifically among participants trained in ACT, assessing whether the anxiety-provoking situation itself could be modified was often described as a preliminary step, leading to action when change was possible: *“In the end, if you can change a situation that’s making you suffer, why would you just stay stuck in it?” (P21)*.

#### Cultivating and sustaining acceptance through practice

> *“I know how it works, in theory. But between knowing and actually doing it, there’s still some work to be done.” (P3)*

Acceptance emerged as a complex process that requires initial learning and ongoing practice and must be enacted rather than merely understood. Theoretical explanations alone were described as insufficient: *“Rationally, you get it pretty quickly—what it is and what you’re supposed to do—but actually doing it… so many times I caught myself thinking, ‘Well yeah, just accept it,’ and actually I was anything but accepting” (P26).* Regular practice was described as central to developing acceptance, which with time and training became more accessible and more natural. “*For me, it’s pretty easy now, yeah… It’s through meditation, through practice. Little by little, with practice, it just comes naturally” (P14)*. Practicing acceptance was reported to support day-to-day emotion regulation: *“I can feel it. I can feel when anxiety is coming. I feel the tension in my body, the heart racing. I’ve developed a stronger bodily awareness, and that awareness allows for self-regulation” (P15)*. Participants also noted that lapses in practice made acceptance harder to access and less effective: “*I really think you have to keep practicing regularly, otherwise it kind of fades, actually” (P5)*. For some participants with extensive meditation practice, acceptance was experienced not as a separate strategy mobilized only in response to difficulty, but as an ongoing stance of openness: *“It’s not like I’m living my life being anxious without noticing and then suddenly trying to open up. It’s actually more the other way around” (P4)*.

#### Ambiguity and misuses of acceptance

> *“I really have to remind myself: this is not about accepting the situation.” (P9)*

Acceptance emerged as a complex, often elusive concept, partly because the term can carry negative connotations and evoke resignation or passivity. Even experienced participants reported ongoing effort to keep in mind that it refers to internal experience rather than the external situation. Even when this distinction was understood conceptually, it was not always apply in practice, and inconsistencies remained *“The situation… Of course there are lots of things we can’t accept. The point is how we make space for what we feel instead of going into a fight with it” (P26).* Acceptance may be misdirected toward tolerating the situation itself—even when perceived as unfair—and used to reduce anxiety, rather than to alter one’s relationship to internal experience. Some participants also described pseudo-acceptance that functioned as avoidance, including shifting attention away from internal experience *“directing attention toward things that are either soothing or just elsewhere” (P2*), using self-reassurance to suppress anxiety “*telling myself there’s no reason to be anxious, that everything will be fine” (P3*), or dismissing emotions “*anger is pointless,” (P19)*. Others framed it as a marker of success in fighting anxiety, which runs counter to its core principle: “*Because there’s also a sense of satisfaction, I think, a kind of pride, in realizing that I finally managed to fight something that had been really disabling for a long time” (P3)*.

### Axis 2 –Acceptance processes

#### Observing through the body

> *“Because I was actually in contact with my senses, I didn’t get carried away by my thoughts anymore.” (P2)*

Practicing acceptance involves reconnecting *“with the body and sensations” (P15),* which helps individuals disengage from repetitive, intrusive thinking and reconnect with the present moment: *“There’s no good or bad thing. Just you, here.” (P12)*. This involves observing bodily sensations by intentionally focusing attention on them. The body or the breath can be used as anchors to bring attention back to sensory experience: “*An anchor means it brings attention back to the center.” (P15)*. This process enhances bodily awareness: “*If you asked me what I feel, I could describe everything, like doing a body scan. So there’s already a strong awareness of the body.” (P6)*, and for some, it was like discovering their body for the first time: “*There was something under the iceberg that I’d never seen before—it was my body. I’d never really seen it.” (P1)*. Participants also pointed to an ambiguity in the role of breathing. It could be used as a soothing technique to reduce anxiety, which is not acceptance per se, insofar as acceptance involves staying open to the experience itself, yet downregulation sometimes served as a preliminary step that facilitates entry into the acceptance process “*For me, I use breathing when it’s hard to open up. I breathe because it allows me to at least bring things down a notch, to take a step back.” (P12*). A similar issue arises with focusing on external senses (sight, hearing, smell, taste, and touch), which can foster mindful awareness but does not necessarily constitute acceptance of anxiety, as it may not involve openness to the anxious experience itself: “*I think that if we stay too much with the breath, we’re focusing on it and eventually it passes, but we haven’t really looked into it. We haven’t explored it deeply.” (P12)*. In addition, participants with an ACT-only framework tended to focus less on bodily sensations than those primarily trained in MBIs, tending toward broader observation of general affective states and thoughts rather than systematically engaging in body-based observation. Direct attention to thoughts tended to reactivate cognitive elaboration (e.g., rationalizing or negotiating), reducing the grounding function of bodily sensations and extending engagement with anxiety.

#### « It’s there » Becoming aware of and identifying the present experience

> *“Maybe it’s when it gets named—when you can put a word on it— that it stops feeling dangerous. I’m no longer under threat, I’m just uncomfortable.” (P18)*

Observation first helped participants notice and acknowledge what was present, an essential step in the acceptance process “*It’s really about realizing that it’s there.” (P21)*. Attending to experience also enabled them to detect the discomfort and identify it “*Sometimes it’s hard to put a name on what we’re feeling, so listening to physical or behavioral reactions helps—it’s easier to spot those than irritation or nervousness.” (P22)* and to name it as anxiety, which appeared to reduce its perceived threat, allowing it to be acknowledged rather than feared and making it easier to attend to its informational value: “*I learned to identify what wasn’t necessarily good for me, but also to see that it carried a message. And then, you know, behind it there’s anger, often sadness, sometimes disgust—there’s this whole mix of emotions.” (P2)*.

#### « It’s OK » Validating and normalizing

> *“It’s telling yourself, actually, it’s OK, we’re human. (…) It’s how we know that something in our environment needs to change. So first, it’s about making peace with the fact that it’s OK to feel anxious.” (P4)*

Participants described a process of validating emotions, most often already identified as anxiety. Validation involved acknowledging the emotion as present and recognizing it as understandable given the situation: “*Basically, what this taught me is that I had the right to feel anxious.” (P10)*. They allowed themselves the experience to be felt: “*I can—I can feel this, in the sense that it’s not going to hurt me, it’s not going to destroy me, it’s not dangerous.” (P18)*. Anxiety was also reframed as a normal and universal aspect of human experience: *“It’s part of who we are, all emotions, whatever they are—it’s human. It’s not… well, it’s normal.” (P3)*. For some participants, this shift went further, with anxiety coming to be understood as meaningful information, an internal signal pointing to personal needs, rather than as a source of shame: *“At first it’s hard because you experience it (anxiety, ed.) as vulnerability, but I don’t anymore. Now I see it as a strength—something that helps you learn about yourself and how to live with it.” (P3)*.

#### Non-reacting

> *“This situation—I can just contemplate it, really. Without talking, without trying to act, without judging it. Just contemplate it.” (P4)*

Observation involved adopting what participants described as an observer stance (i.e., stepping back from the experience, creating distance, and relating to it from the outside): *“I feel like I’m taking a step back and having this kind of inner gaze, or witness, that looks in a more neutral way. Getting out of that emotional fog so things can settle.” (P10)*. This stance allowed to notice habitual thought patterns and automatic reactions: “*That’s when I really saw the scenarios playing out. So then it’s like, stop—I slow things down.” (P 26)* while deliberately refraining from reacting, involving recognizing the struggle as resistance to internal experiences: “*I don’t want to go through this right now! I don’t want to experience this!” (P26)* and letting go of effort to control anxiety by changing it, pushing it away, or judging: “*You step out of judgment, out of that urge—more or less—to make things happen this way or that way, to control it.” (P12).* Among ACT-trained participants, this phase also involved assessing whether action on the triggering situation was possible: *“You have to be in that space of: OK, what’s happening inside me? And then ask yourself: Can I act here? Or can’t I?” (P21)*.

#### Staying with the experience

> *“And then opening to it—well, letting it be there. Like: OK, you’re here, go on then, take up the space you need, mess me around in my body if you want, too bad—and that’s it. And it lasts as long as it lasts.” (P12)*

Participants highlighted that acceptance requires staying with anxiety, even when unpleasant, without reinforcing or intensifying it: “*OK, you have a right to exist. I don’t really want you here, but I’ll let you do your thing.” (P12)*. This required a clear awareness of personal limits to remain within tolerable distress: “*What’s the balance between my desire to move toward the unknown, toward discovery, and at the same time not being in too much discomfort—definitely not in danger.” (P24)*, while continuing to live and act, rather than becoming paralyzed by anxiety. To stay with experience longer, participants described deliberately extending observation: *“Keeping yourself occupied with an exploration that actually allows you to stay and observe.” (P14)*.

### Axis 3 – Acceptance-related outcomes

#### Transformation of the experience

> *“I feel relieved—I tell myself I don’t need to fight it, I don’t need to go against it.” (P1)*

Across participants, acceptance was associated with feelings of relief, calming “*it settles down” (P8)* and a sense of lightness, even when unpleasant sensations persisted: “*Feeling calm while still being anxious.” (P23)*. Participants described acceptance as reducing rumination by redirecting attention away from intrusive thoughts and toward bodily sensations: “*It helps me not get stuck in loops of rumination.” (P15).* As repetitive thinking diminished, acceptance was said to free up “mental space,” allowing participants to regain a sense of agency rather than remaining locked into automatic, anxiety-driven reactions: “*I was no longer guided by my anxiety, always like, ‘Quick! I have to deal with this, deal with that.’” (P5)*, supporting a more open, creative stance toward challenges: “*It becomes easier for me to see things, to come up with ideas about what I could do.” (P18).* Participants also described a broader perspective on experience, with anxiety becoming one element among others rather than totalizing: *“To widen awareness, too—not just what’s unpleasant, but also what’s pleasant… It might be a diffuse discomfort, but it’s pretty rare that everything is tense; there are usually places that feel more relaxed, more settled, and you can come back to that as well.” (P6)*.

#### Transformation of the self-perception

> *“I’ve learned to better understand how I react to certain events, to make sense of those reactions, and as a result to regulate some of my states more effectively in everyday life.” (P 7)*

Engaging in acceptance fostered deeper self-understanding among participants, including greater awareness of habitual patterns, emotional responses, and personal tendencies: *“I’ve learned more about what really matters to me, about my blind spots, my tendencies, my impulsivity, and also my resources.” (P18)*. This growing self-knowledge was often accompanied by greater compassion and self-kindness, described as *“treating myself more kindly” (P5)* and increased openness and empathy toward others, with less judgement: *“It’s really this awareness that I’m just like everyone else, that I have plenty of flaws too, which gives me the ability to open up to others now.” (P10)*. Some participants described a broader sense of connection or belonging, extending to others or to the world more generally. Acceptance was further associated with calm, inner steadiness, and greater self-confidence *“I also feel more grounded, and that gives me this sense of confidence.” (P7),* as well as feeling freer to be themselves *“I’ve learned to hide less, to be more authentic” (P2)* and more able to experience positive emotions alongside anxiety.

### Grounded Theory

Engaging in the acceptance process entails a deliberate and proactive decision, which initially involves stepping back from anxiety and, when necessary, downregulating emotional arousal to allow entry into the process. In some contexts, individuals may also decide not to engage in acceptance if it is perceived as unsuitable in a particular context, instead opting for an alternative emotion regulation strategy. Several factors appear to facilitate and strengthen the acceptance process, including support provided by MABIs frameworks, psychoeducation, engagement with meaning and values, experiential practice, regular training, and self-compassion. Our grounded theory indicates that anxiety acceptance process involves five interconnected stages: (Stage 1) observing through the body, (Stage 2) identifying, (Stage 3) validating and normalizing, (Stage 4) not reacting, and (Stage 5) staying with the experience. Moreover, the findings indicate that acceptance transforms the experience of anxiety. While anxiety does not necessarily disappear, it is experienced differently, with reduced struggle and a shift in perspective. This transformation is associated with enhanced agency, improved self-knowledge and self-confidence, and more satisfying interpersonal relationships. Overall, this model highlights the stages underlying the experience of acceptance, its perceived benefits, and the central role of active mobilization to engage in this process (Fig 1).

**Fig 1.** Grounded theory of anxiety acceptance.

## Discussion

This qualitative study identified five stages characterizing participants’ experiences of acceptance, along with factors that moderate their deployment and the effects of acceptance on individuals and their relationship to anxious experiences. Taken together, these analyses led to the development of a comprehensive explanatory model of acceptance in the context of anxiety, encompassing mobilization, enactment, and subsequent transformations. Focusing on the implementation of acceptance, the model highlighted the mechanisms and processes involved. Participants’ phenomenological accounts described combinations of stages, including (Stage 1) observing through the body, (Stage 2) “It’s there”: becoming aware of and identifying the present experience, (Stage 3) “It’s OK”: validating and normalizing, (Stage 4) non reacting, and (Stage 5) staying with experience. These stages can be linked to several theoretical constructs from the mindfulness and emotion regulation literature and our identification of the underlying mechanisms and processes yielded a detailed descriptive model of the acceptance process (fig 2).

**Fig 2.** Modeling the stages of acceptance and their correspondence with subprocesses.

### Stage 1: Observing through the body

This stage involves attending to anxiety through bodily sensations by directing attention either toward anxiety-related somatic cues or toward a bodily anchor (e.g., the breath or sensory input), thereby facilitating disengagement from intrusive anxious thoughts. This stage supports the development of a more acurate perception of internal bodily sensations. These findings align with the concept of interoception within the emotion regulation literature (58), as well as to bodily awareness (29) and attentional regulation (21) within the mindfulness framework. Greater sensitivity to internal bodily sensations is associated with improved abilities to recognize and understand emotions (58) and more effective emotion regulation (59–62). Thus, observing anxiety through the body appears to initiate emotion regulation processes.

### Stage 2: Noticing and identifying the present experience

This stage consists of recognizing and labeling the experience as anxiety. These findings align with concepts of emotional identification within emotional competence frameworks (63), emotion labeling (26,64), and the first level of emotional validation (17). Identifying or labeling emotions facilitates a clearer understanding of perceived emotional states and guides regulatory strategy selection, constituting a form of implicit emotion regulation (65) that is effective across multiple domains (66). It has been associated with modulation of emotional responding, including reduced affective intensity (67), effects comparable to cognitive reappraisal (68), and psychological and physical health benefits (69). These effects may be explained by self-reflection or self-distancing (70), uncertainty reduction via categorization (71,72), and increased psychological distance through abstraction (65) and verbal encoding, facilitating the downregulation of amygdala alarm responses (73). Overall, labeling may foster a decentering-like distancing from emotion, supporting a more differentiated, less fused relationship to experience. Consistent with this account, participants reported that naming anxiety reduced its burden and anticipatory concern: noticing discomfort established its presence, whereas identifying it as anxiety made it less threatening and more tolerable, reducing inadvertent maintenance and supporting regulation.

### Stage 3: Validating and normalizing

This stage involves recognizing anxiety as legitimate given the context, normalizing it as a universal phenomenon inherent to the human condition, and viewing it as a potentially adaptive signal. Our findings converge with models of emotional validation (74), self-compassion (51), and with the emotional competence of understanding emotions (63), which considers emotions as adaptive responses to context (75,76) with an informative function regarding individual needs (77). Emotional validation refers to acknowledging the existence of an emotion and legitimizing it, which can reduce distress and defensive reactions and support tolerance and adaptive regulation strategies (78). Consistent with this, self-compassion (self-kindness and common humanity) facilitates the learning and use of emotion regulation skills (e.g., 50,79). In our model, these processes support access to acceptance, promotes emotion regulation, reduces global reactivity and enhance distress tolerance.

### Stage 4: Non-reacting

The ability to observe internal experiences (i.e., thoughts, emotions, sensations) requires stepping back from anxiety to notice it without reacting (i.e., judging, struggling against, rejecting, reassurance seeking or resisting). These findings align with decentering (80), the FFMQ dimensions of nonreactivity and nonjudgement (26), and response prevention in exposure-based therapies (30,81,82). Decentering refers to stepping back from experience and viewing it from an observer-like stance (80), viewing internal experiences as transient and objective events rather than accurate self-representations (83) and comprises meta-awareness (i.e., awareness of subjective experience as a process), disidentification from internal experience (i.e., viewing internal states as separate from the self), and reduced reactivity to internal content. It underlies related constructs across models, including cognitive defusion and self-as-context in ACT (14,39), metacognitive awareness in MBCT (84), the metacognitive mode (85), and reperceiving (23). Decentering is a proposed mechanism of change in anxiety treatments (86,87) and a key mechanism underlying emotion regulation in MABIs (88). Consistent with prior work (12), our findings place decentering as a subprocess within acceptance. Similarly, response prevention inhibits habitual, avoidance-reinforced responses and promotes habituation and reduced anxiety, closely aligning with Stage 4, which involves a distanced observational stance that inhibits immediate reactivity. It represents an intermediate yet essential skill within acceptance, thereby supporting more effective emotion regulation.

### Stage 5: Staying with experience

This stage involves active engagement with anxiety by staying with it and allowing discomfort to unfold without attempts to alter it, functioning as interoceptive exposure, which may increase distress tolerance over time. These findings are consistent with exposure and response prevention models (ERP, 30) and distress tolerance frameworks (89). Exposure involves remaining in contact with difficult emotional experiences without avoidance to reduce their impact through habituation, leading to the extinction of fear responses (22) and interoceptive exposure may reduce emotional reactivity (90). Distress tolerance, the ability to stay in contact with unpleasant emotional states without attempting to alter or escape them (89), rather than fear reduction, has been proposed as a primary target of exposure-based interventions (82) and is associated with less reliance on maladaptive strategies (e.g., suppression, avoidance, or rumination) and greater habitual use of adaptive strategies (9,93). Accordingly, stage 5 may function as exposure to anxiety-related discomfort, promoting distress tolerance and an approach-oriented stance that supports acceptance.

### Toward acceptance as a multidimensional and dynamic process-based model

After examining the processes underlying acceptance separately, we then considered how they integrate within our overall model and how this model relates to existing theoretical frameworks as summarized in Table 2. Several processes overlap with mechanisms embedded in broader mindfulness models (e.g., 21,26,29), ACT (14,39), or models rooted from cognitive-behavioral therapies, including exposure with response prevention (30). However, these frameworks capture our processes only partially, leaving several key dimensions of our model less fully addressed. In our empirically grounded model (see Figure 2), acceptance emerges as a set of interrelated processes and active engagement with emotional experience, supporting its conceptualization as a multidimensional, complex and dynamic phenomenon unfolding over time. Identified as the active ingredient of MBIs (18), acceptance can thus be considered a process of change (2–4), reflecting a dynamic set of cognitive, emotional, and behavioral operations through which individuals respond to internal experience in ways that can transform it. Moreover, our model suggests that each stage of the acceptance process involves emotion regulation: attending to bodily sensations engages regulation by shifting attentional focus and modulating arousal; labeling anxiety contributes to implicit regulation by clarifying the emotional experience and reduces its perceived threat; validation alleviates secondary distress linked to self-judgment and resistance; non-reactivity limits maladaptive regulatory responses (e.g., avoidance, suppression, struggle); and staying with the experience promotes distress tolerance over time. Accordingly, acceptance functions as a dynamic, multi-process mode of emotion regulation involving decentering, self-compassion, emotion labeling, emotional validation and interoceptive exposure.

**Table 2.**
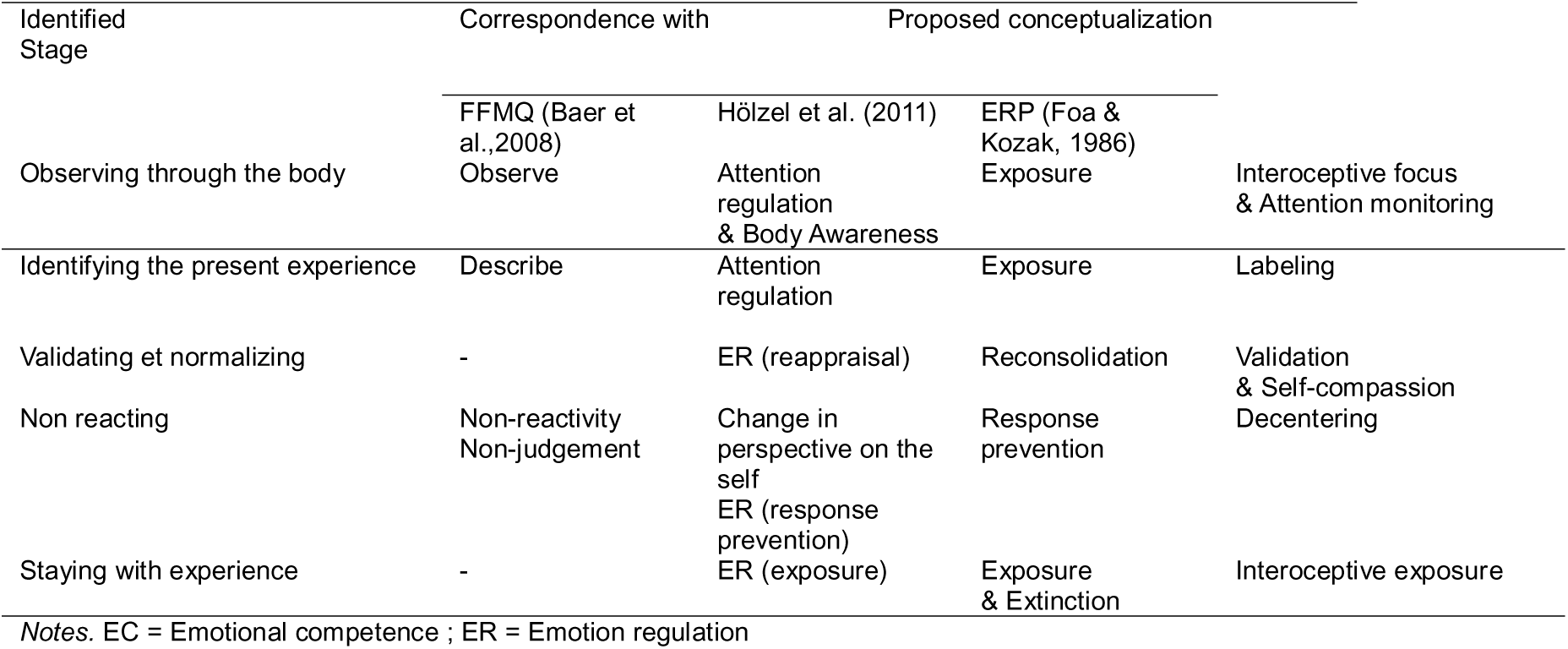
Correspondence of Observed Acceptance Processes With Existing Concepts and Theoretical Models.

### Mobilizing toward acceptance as a critical step

Our findings highlight the necessity of a deliberate and proactive decision to initiate the process of acceptance. However, anxiety can be experienced as paralyzing, most often due to its intensity, which may hinder access to acceptance by limiting the mental availability required to engage in it. This difficulty is consistent with models of attentional bias and attentional control, which posit that anxiety captures attention toward threatening stimuli and impairs executive functions, thereby making disengagement more difficult (94–97). Moreover, attentional control has been shown to be closely related to emotion regulation, suggesting that the effectiveness of emotion regulation partly depends on this higher-order mechanism (98). It therefore seems reasonable that intense anxiety may impede disengagement and limit effective mobilization toward acceptance. Mindfulness and more specifically the nonreactivity facet of the FFMQ (26), which has been considered a proxy for acceptance in some studies (e.g., 27,99), has been shown to facilitate disengagement from emotional stimuli (100). In addition, mindfulness training explicitly targets attentional control and has been shown to enhance it (101). Orienting toward acceptance may also foster a sense of openness that broadens the scope of awareness (102) and frees attentional resources to notice and savor positive experiences in the present moment (103). Consistent with this, engagement in acceptance appeared easier among participants with longer experience and regular training. This initial mobilization may be understood as a brief pause, or as an emerging form of decentering, described by participants as a “step aside” requiring a conscious effort to disengage from the sense of urgency induced by anxiety. The stages of our model may thus be simultaneous rather than strictly sequential. This perspective aligns with MABI approaches conceptualizing acceptance as an active and voluntary stance (14), involving the choice to face aversive internal experiences rather than avoid them. The temporal ordering of decentering and acceptance remains unclear and may depend on emotional activation: in low-arousal, decentering may foster acceptance, whereas under high emotional activation, acceptance may enable decentering (104). Nonetheless, orienting toward acceptance may broaden awareness and release attentional resources (102,103), consistent with participants’ descriptions of “freeing up mental space”, suggesting that adopting an accepting stance is already a first step that provides the attentional availability required to engage in the processes that comprise acceptance.

### Factors influencing acceptance and the core role of self-compassion

Several factors were identified as influencing both the mobilization toward acceptance and its implementation, and most notably anxiety intensity Acceptance was generally feasible at low to moderate levels, whereas intense anxiety often required preliminary downregulation, or the use of other strategies, before acceptance could be engaged. Other factors also appeared to shape the capacity for acceptance, including contextual resources such as available time, energy, the possibility of being alone, and external supports, particularly close others who may serve as sources of compassion and validation. In addition, MABIs provide a supportive context for learning and practicing acceptance through formal and informal meditation, and greater practice duration, frequency and regularity appear to facilitate mobilization toward acceptance by fostering more automatic interoceptive attentional focus. This pattern is consistent with previous findings indicating that disengagement-based or perseveration-based emotion regulation strategies tend to be more automatic under conditions of high emotional activation (9). As a adaptive engagement strategy, acceptance may therefore be less spontaneous and require an initial learning phase to be effectively enacted (105–107), with at least one to two weeks of training to observe meaningful improvement (108) or roughly 5–10 hours of mindfulness training to observe differentiated mechanistic effects (19). These findings suggest that learning and practice are prerequisites for effective acceptance. Brief acceptance-based trainings may benefit only individuals with prior experience (109) and greater formal practice predicting stronger MBI benefits (110). In addition, experienced meditators show lower anxiety than novices (106), and more frequent mindfulness practice has been associated with greater well-being regardless of experience level (111). Taken together, learning and regular practice appear to moderate acceptance effectiveness.

Self-compassion was another key factor, facilitating mobilization, supporting practice, and shaping the acceptance process across multiple stages of our model, particularly Stage 3 (validation and normalization), where it supports self-kindness and common humanity, and Stage 4 (non-reacting), where it helps individuals refrain from judging or criticizing internal experience. For some participants, self-compassion marked a turning point, enabling a fuller, more experiential acceptance rather than continued struggle. Overall, self-compassion may both moderate acceptance (i.e., facilitating it when higher) and mediate it as a process that enables access to and implementation of acceptance, thereby contributing to acceptance’s effectiveness and amplifying its impact. Notably, MBIs explicitly cultivate self-compassion: MBSR increases self-compassion (112–114), and self-compassion may mediate change within MABIs (Baer, Lykins, et al., 2012). More broadly, self-compassion is associated with more adaptive emotion regulation (49,50). These findings are consistent with our results suggesting that self-compassion may mark a pivotal shift in acceptance practice, facilitating a transition from an intellectual understanding to experiential engagement. Fully enacting acceptance may therefore be difficult, if not impossible, without mobilizing self-compassion.

### Changes associated with acceptance

The third axis focuses on the effects attributed to accepting anxiety, on emotional experience and self-relation, consistent with MABI models. First, accepting anxiety appeared to reduce feelings of overwhelm, particularly by reducing aversive repetitive thoughts, often described as among the most distressing aspects of anxiety. These findings are consistent with prior research showing that MABIs reduce the impact of intrusive aversive thoughts (115) by fostering greater distance from them (16). In this regard, reconnecting with the body through observation and anchoring in bodily sensations, seems to play a key role by shifting attentional focus away from ruminative thought. Acceptance changes one’s relationship to anxiety without necessarily reducing its inherent tension, underscoring the distinctive goal of this strategy rather than direct downregulation, and highlighting that it should not be evaluated solely by symptom reduction but rather by distinct criteria. Nonetheless, accepting anxiety can bring relief by ending the struggle, reducing cognitive hyperactivation, clarifying the present experience, and broadening awareness so anxiety becomes one element among others rather than the dominant focus. Acceptance also modifies the relationship to action in the context of anxiety by creating space between the emotional experience and behavioral responses, enabling more intentional, flexible, values-consistent choices rather than anxiety-driven behavior. This shift supports deliberate action and a renewed sense of agency. Beyond these immediate effects, our findings align with prior work identifying acceptance as protective against emotional distress and conducive to personal growth. Acceptance promotes enhanced self-observation and self-understanding, allowing individuals to identify their patterns, better comprehend their reactions, and recognize their personal resources. Participants also reported increased self-confidence, grounded in improved self-knowledge and a perceived ability to accept their emotions, suggesting a strengthening of self-esteem. This is consistent with evidence indicating a positive impact of mindfulness on self-esteem (116,117), as well as the development of a more compassionate and benevolent stance toward oneself. Acceptance was also associated with improved interpersonal functioning, with participants reporting greater understanding and compassion toward others, consistent with evidence that mindfulness enhances prosocial behaviors and compassion (e.g., 118). By fostering openness to anxiety, validation, and a sense of shared humanity, acceptance may increase awareness of the universality of suffering which promotes greater empathic concern and altruism (119), suggesting that acceptance may support the development of empathy, defined as the capacity to perceive and understand others’ internal states (120). Finally, acceptance appears to create space for positive emotions beyond anxiety, in line with the Mindfulness-to-Meaning Theory (121) which posits that mindfulness facilitates cognitive and emotional reappraisal under stress, allowing individuals to derive positive meaning from lived experiences. Taken together, these changes suggest pathways through which acceptance contributes to emotion regulation.

### Limitations and directions for future research

The present study has several limitations. From a methodological perspective, participant accounts may have been influenced by prior theoretical knowledge acquired through MABIs, rather than reflecting only lived experience. However, precautions were taken to promote phenomenologically grounded accounts, including the use of targeted follow-up questions, the solicitation of concrete and context-specific experiential narratives, and the exclusion of overly theoretical discourse during data analysis. Although theoretical saturation was achieved in accordance with Grounded Theory principles which prioritize saturation over quantitative representativeness, the sample may not capture more singular or atypical experiences, limiting generalizability without further validation. In addition, the explicit focus on acceptance may have contributed to an overestimation of its role, even though participants reported using other regulation strategies and our aim was to characterize acceptance rather than estimate its relative importance. Furthermore, the coding process inevitably reduces experiential complexity despite an inductive and reflexive approach intended to preserve data richness; thus, our model captures a general dynamic rather than the full singularity of each case, and further research could refine the dimensions identified here. Finally, potential biases (i.e., social desirability, self-selection, positivity bias, retrospective memory reconstruction) cannot be ruled out and the analysis is also situated within a specific conceptual framework and remains dependent on the researcher’s interpretive stance. Accordingly, the findings reflect subjective experience and cannot be direct linked to objective mental health indicators, underscoring the need for complementary research, including quantitative studies, although existing evidence broadly supports our conclusions.

Despite these limitations, the present study contributes to the understanding of acceptance and highlights several promising directions for future research. The proposed process-based model would benefit from empirical validation, particularly through longitudinal quantitative studies examining the sequencing, stability, and necessity of the identified stages and subprocesses, as well as their variation across different forms of anxiety (e.g., panic attacks, generalized anxiety) or other emotions (e.g., anger, shame). Developing multidimensional measures grounded in these processes also appears warranted. Finally, the facilitating role of self-compassion merits further investigation, including its potential moderating and mediating functions and its targeted integration into interventions. Together, these lines of inquiry may help refine the conceptualization of acceptance and support more precise and personalized clinical interventions.

### Practical implications and clinical applications

The findings of this study have direct clinical relevance by offering concrete guidance to refine how acceptance is taught and learned within MABIs and to inform tailored interventions. Because acceptance can be difficult to articulate or grasp verbally (122), patients may misunderstand what to do in practice; using terms such as openness or welcoming, while emphasizing that acceptance targets internal experience rather than external reality, may reduce conceptual confusion. The structured model proposed in this study provides a didactic framework for progressive, targeted learning of acceptance. By identifying which subprocesses are most challenging for a given individual, clinicians can implement focused, idiographic, process-based work (4), thereby enhancing the effectiveness of therapeutic or preventive interventions. Our results also underscore self-compassion as a key facilitator, supporting its explicit integration within MABIs.

## Conclusion

The present study led to the development of a comprehensive process-based model of accepting anxiety, conceptualizing acceptance as a dynamic set of interdependent subprocesses and providing a concrete operationalization of how it emerges and unfolds during anxiety, including key individual and contextual factors that shape its unfolding. We also identified major obstacles to implementation, particularly difficulties mobilizing toward acceptance under high perceived anxiety intensity. Our findings also highlight the central role of self-compassion in enabling this process and show that acceptance is associated with benefits extending beyond shifts in anxiety experience to broader changes in self-relation, underscoring its relevance for emotion regulation and well-being.

Finally, the study clarifies persistent ambiguities around acceptance by documenting common inconsistencies in meaning and practice. Together, these results refine the conceptualization of acceptance and inform process-based research and clinical work by identifying actionable subprocess targets, guiding therapeutic interventions, anticipating obstacles, and supporting individualized intervention.

## Data Availability

Full interview transcripts are not publicly available due to ethical and confidentiality restrictions, as they contain sensitive, potentially identifying contextual information. Participants consented to scientific use of pseudonymized data and to the publication of anonymized quotations, but not to public release of full transcripts. This was approved by the Ethics Committee of University Paris 8 ( protocol #CE-P8-2021-07). De-identified excerpts supporting the findings may be shared with qualified researchers upon reasonable request, subject to ethical review and a data sharing agreement. Requests: secondary contact

## Notes

### Competing Interest Statement

The authors have declared no competing interest.

### Funding Statement

The author(s) received no specific funding for this work.

### Author Declarations

The Ethics Committee of Universite Paris 8 Vincennes Saint Denis gave ethical approval for this work.

